# Developing an online cannabis store paradigm for Cannabis Regulatory Science

**DOI:** 10.1101/2025.10.20.25338387

**Authors:** J.T. Borodovsky, S. Bairaboina, C.A. Struble, S.M. Preum, J.D. Sargent, J.A. Emond, A.J. Budney

## Abstract

**Background:** The legal cannabis industry is using online point-of-sale marketing, but few tools exist for evaluating how this marketing influences consumer behavior. To address this gap, we developed the Platform for Online Evaluation of Marijuana Marketing and Sales (POEMMS), a realistic, customizable online cannabis store. This study tested the feasibility, usability, and ecological validity of POEMMS.

**Methods:** U.S. adults aged 18+ were recruited via social media advertisements. Participants engaged in a simulated shopping task using POEMMS and then completed a post-shopping survey. Participants were instructed to imagine having no cannabis and to buy products as they would in real life. Survey measures included typical real-world cannabis spending and 0–10 Likert ratings of the store’s usability and realism. Spearman correlations tested associations between in-store and self-reported spending.

**Results:** N=678 completed the shopping task. Approximately 58% used cannabis daily. Median total spending was $100. Shopping task spending and self-reported spending for flower, open concentrates, cartridges, and edibles were strongly correlated (rs = 0.74, 0.74, 0.67, 0.62, respectively; all p<0.001). Participants rated the store as easy to navigate (Median=10) and product characteristics easy to find (Median=10). Perceived realism was also high across product names, types, potency, and descriptions (Median=8–10).

**Conclusions:** POEMMS elicited behaviorally and perceptually valid responses in a large national sample of cannabis users. Findings demonstrate the platform’s utility for Cannabis Regulatory Science by allowing controlled evaluation of online marketing effects. Future applications of POEMMS could generate evidence to inform cannabis marketing policy and regulation.

## 1. INTRODUCTION

U.S. legal cannabis is a multi-billion-dollar industry that has been described as “a natural laboratory for marketing strategy research”(Olsen and Smith, 2019). In the wake of the COVID-19 pandemic, the industry has made a substantial shift to online sales and advertising (Israel, 2021; Jackson, 2020a, 2020b; Rup et al., 2020). However, regulatory frameworks have not kept pace with this transition, and thus, online cannabis retailers often operate with minimal oversight (Berg et al., 2023a). Consequently, cannabis store websites frequently employ aggressive point-of-sale (POS) marketing strategies that combine misleading pharmaceutical-style health claims and recreational lifestyle branding while omitting or downplaying adverse consequences and health warnings (Borodovsky et al., 2021; Borodovsky and Budney, 2018; Hoeper et al., 2022; Rhee and Jung, 2023). Given that cannabis consumers consider multiple product attributes beyond just price or potency (e.g., perceived quality, strain, packaging, recommendations) marketing strategies could shape purchase decisions and increase health risks among vulnerable subgroups such as young adults and those with mental health problems (Borodovsky and Budney, 2018; Donnan et al., 2022a, 2022b, 2024; Shi et al., 2019; Trangenstein et al., 2021; Zhu et al., 2021).

Despite rapid expansion of online cannabis retail, research on POS marketing within online stores remains limited, which leaves policymakers without the data needed to inform regulation. Much of the existing research on online cannabis marketing has focused on social media rather than the marketing used *within* online stores and/or use observational methods that do not allow for direct testing of marketing effects on purchase decisions (Duan et al., 2023; Marinello et al., 2024; Rup et al., 2020; Whitehill et al., 2020). Some studies have examined cannabis-related health claims, branding, and warning labels using survey-based experiments (Donnan et al., 2023; Goodman et al., 2019; Kowitt et al., 2022; Mutti-Packer et al., 2018), which limits ecological validity. Furthermore, studies that have investigated cannabis POS marketing have focused on brick-and-mortar stores (Berg et al., 2023b; Cao et al., 2020), rather than the store websites. Thus, there is a need for a research system that enables controlled experimental testing of online POS marketing strategies in a realistic cannabis shopping environment.

To address this gap, we developed the Platform for Online Evaluation of Marijuana Marketing and Sales (POEMMS)—a realistic, customizable online cannabis shopping platform. POEMMS allows researchers to control product attributes and display designs, implement various in-store advertising techniques (e.g., customer reviews and product recommendations, discounts, pop-up messaging, etc.), and track purchasing behavior in real time. This study presents pilot results on the feasibility of POEMMS for Cannabis Regulatory Science by evaluating the platform’s usability, realism, and validity.

## 2. METHODS

### 2.1. Platform Development and Structure

POEMMS consists of a front-end online cannabis store and a back-end digital infrastructure for data collection and product stimuli manipulation. The platform was built using WordPress® and WooCommerce®, which are the same systems used by many real online cannabis stores. The back-end infrastructure can track behaviors, including time spent browsing, page clicks, product views, and purchase decisions. For this initial study, the front-end store was populated with realistic cannabis product stimuli (e.g., pictures, names, potency, prices, and descriptions) based on peer-reviewed literature, expert consultation, and industry sources (e.g., Dutchie®, Wikileaf®, Leafly®). The initial version of the store was programmed to offer four popular product categories: flower/bud, open concentrates (e.g., wax, shatter), prefilled concentrate vaporizer cartridges, and edibles (Figure 1). Each product category contained six individual products that can vary on factors believed to affect purchase decisions, e.g., THC/CBD percentages, prices, and names. Participants can click on any of the six product variations to view a detailed individual product page displaying other factors that might affect purchase decisions (e.g., strain characteristics, expected effects) as well as an “add to cart” button (Figure 2).

**Figure 1.**
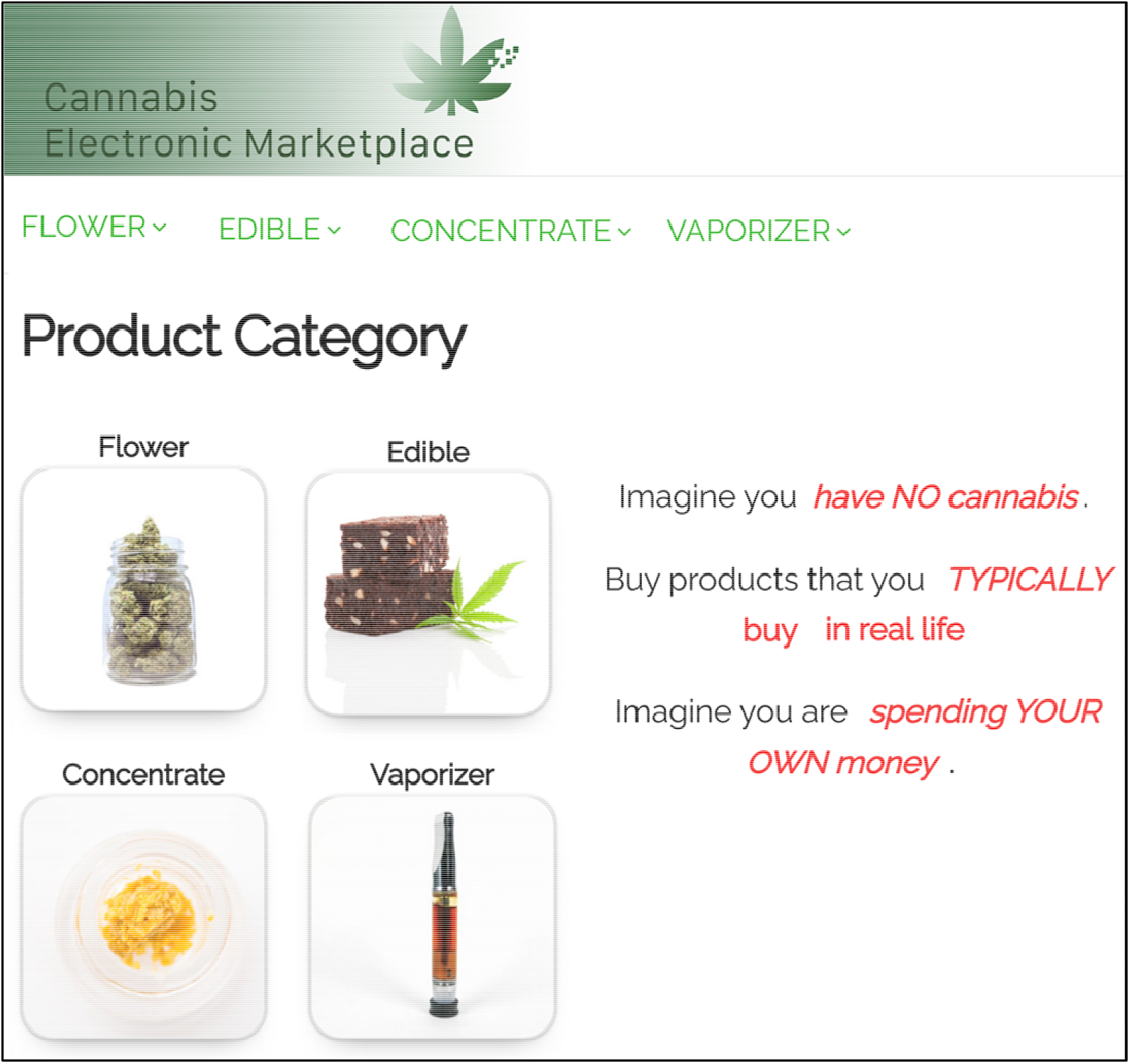
Screenshot of the homepage of the POEMMS online dispensary interface. POEMMS was built using WordPress® and WooCommerce® to mimic style and design elements of commercial cannabis retail websites. Participants selected from the four product categories (flower, edibles, open concentrates, and vaporizer cartridges). The standardized task instructions (red text), were displayed on all store pages.

**Figure 2.**
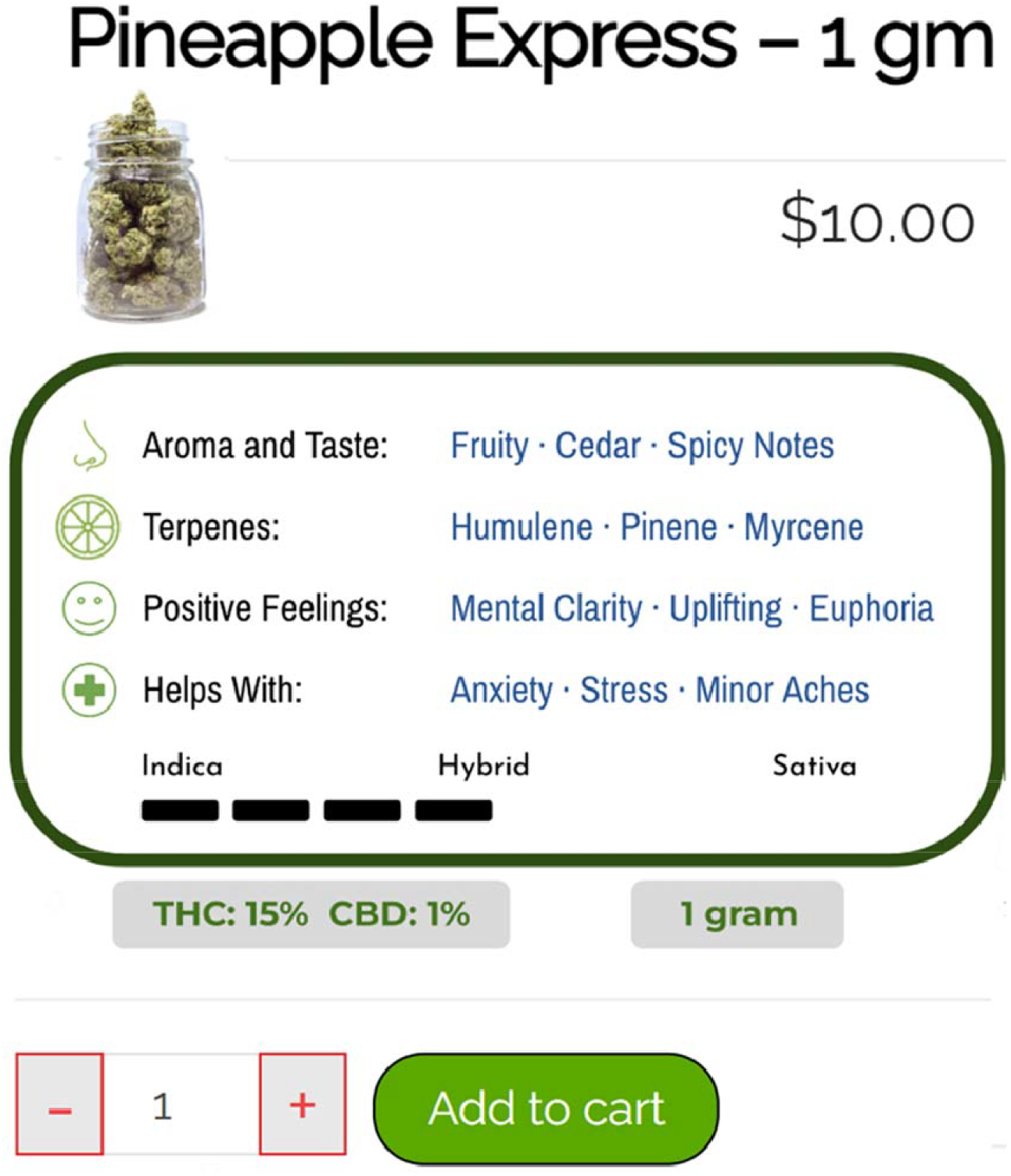
Screenshot of a POEMMS individual product page displaying product-specific information and purchase controls. Each product page included price, THC/CBD content, strain characteristics (e.g., aroma, terpenes, reported effects), indica-sativa profile, and an “add to cart” button. This content was standardized across products and designed to reflect common marketing elements used by commercial cannabis retailers.

### 2.2. Participant Recruitment and Eligibility

Participants were recruited (9/1/2023 to 10/31/2023) via Meta® advertisements displaying a cannabis leaf and targeting U.S. adults aged 18+ with cannabis-related interests (e.g., Bob Marley)(Borodovsky et al., 2018). Participants were not required to have previously purchased cannabis online to participate. Advertisements directed individuals to the study landing page, where they provided consent and were redirected to the store. IP addresses were tracked to prevent duplicate responses. No participation compensation was provided. The study was approved by the Dartmouth Committee for the Protection of Human Subjects.

### 2.3. Shopping Task and Instructions

Participants were instructed to (1) imagine they had no cannabis, (2) buy products they typically purchase in real life, and (3) shop as if spending their own money.(Aston et al., 2021) These instructions were viewable on every webpage within the store (Figure 1). Shopping was self-paced. Participants also completed a post-shopping survey. The study instructions avoided revealing the primary research questions to mitigate demand characteristics.

### 2.4. Post-Shopping Survey

After shopping, participants completed a post-shopping survey that queried demographics, cannabis use patterns, and typical real-life spending amounts on cannabis flower, prefilled concentrate cartridges, open concentrates, and edibles. Specifically, participants were asked “When you buy cannabis in real life, about how much money do you typically spend on [flower/cartridges/concentrates/edibles]”. Additionally, to assess the usability of the store, participants used a Likert scale from 0 (Very Difficult) to 10 (Very Easy) to rate the difficulty of (1) navigating the store, (2) following the study instructions, and (3) finding relevant product characteristics (e.g., %THC, price). Participants also used a Likert scale from 0 (Very Fake) to 10 (Very Realistic) to rate the realism of the store product names, types, potencies, prices, and descriptions.

### 2.5. Analysis

We used descriptive statistics to summarize (1) demographics, (2) time spent shopping (3) shopping task purchase behaviors, (4) usability ratings, and (5) realism ratings. To assess the reliability of responses, we used Spearman correlations to compare purchase amounts observed in the shopping task to self-reported typical real-life purchase amounts. All analyses were conducted using Python 3.11.

## 3. RESULTS

### 3.1. Sample

N=852 participants consented, N=678 purchased at least one product, and N=579 answered at least one post-shopping survey question. Participant mean age was 36.6 years (SD: 12.9); 52.5% were female, and 80.0% were White. Nearly half (49.0%) reported an annual household income of ≥$50,000, and 37.7% had a bachelor’s degree or higher. More than half (57.9%) reported daily cannabis use in the past 30 days.

### 3.2. Shopping patterns

Participants shopped for a median of 2.58 minutes (IQR: 1.72-3.65 minutes) and viewed individual products for a median of 10 seconds (IQR: 5-16 seconds). Participants purchased a median of 2 (IQR: 2-4) individual products. Median total spending was $100 (IQR: $60-$194) and was highest for flower products (Median: $45, IQR: $15-$105) followed by prefilled cartridges (Median: $40, IQR: $0-$50), open concentrates (Median: $0, IQR: $0-$25), and edibles (Median: $0, IQR: $0-$20). These spending patterns are highly consistent with data from other published studies and industry reports (D’Amico et al., 2020; Gebru et al., 2023; Kepple et al., 2016; Kepple and Freisthler, 2018; McCann, 2024; Olson, 2015; Statista, 2020; Team Headset, 2020). Participants’ spending in the shopping task correlated strongly with their self-reported typical real life cannabis spending for flower (r_s_ = 0.74, p < 0.001), open concentrates (r_s_ = 0.74, p < 0.001), vaporizer cartridges (r_s_ = 0.67, p < 0.001) and edibles (r_s_ = 0.62, p < 0.001). Most participants (91.6%) agreed or strongly agreed that the products they purchased during the shopping task were similar to their usual real-world purchases.

### 3.3. Usability and Realism Ratings

Participants rated the store as easy to navigate (Median: 10, IQR: 8-10), the study instructions as easy to understand (Median: 10, IQR: 10-10), and product characteristics as easy to identify (Median: 10, IQR: 7-10). Realism ratings were high for product names (Median: 10, IQR: 8-10), product types (Median: 10, IQR: 8-10), potency (Median: 8, IQR: 6-10), and descriptions (Median: 9, IQR: 7-10). Realism ratings for prices were more uniformly distributed (Median: 6; IQR: 4-8.5).

## 4. DISCUSSION

This study examined the feasibility of the Platform for Online Evaluation of Marijuana Marketing and Sales (POEMMS) as a tool for studying cannabis purchasing behaviors in an online retail environment. Participants rated the store as highly realistic and easy to use, which suggests that the platform effectively replicates key aspects of the online cannabis shopping experience. Furthermore, POEMMS elicited shopping behaviors that were internally consistent with self-reported real-world cannabis spending, as reflected by strong correlations between in-store purchases and post-shopping survey responses. Observed spending patterns qualitatively mirrored purchase amounts reported in prior cannabis studies and industry data, suggesting that participants’ behavior in the simulated store resembled real-world consumer behavior. Overall, these results support the ecological validity of POEMMS and its potential as a tool for studying cannabis consumer decision-making.

As the online cannabis industry expands, understanding how point-of-sale marketing strategies influence consumer behavior will be increasingly important. POEMMS provides a structured environment for investigating this issue by balancing experimental control with ecological validity. Because the platform is highly customizable, researchers can systematically modify and test the impact of marketing-, pricing-, and policy-relevant variables, such as product descriptions, packaging, labeling, branding, customer reviews, bundled discounts, taxation strategies, warning labels, and health risk messaging. Findings from such studies could be used to understand how cannabis industry and regulatory practices shape consumer behavior and impact public health (Borodovsky et al., 2021).

One area of particular concern is the cannabis industry’s growing use of psychotherapeutic advertising claims (PACs)(Boatwright and Sperry, 2020; Duan et al., 2023; Hoeper et al., 2022; Lau et al., 2021; Luc et al., 2020). PACs contain mental health proxy terms (e.g., ‘happy,’ ‘relaxed,’ ‘uplifted’) and/or explicit treatment claims (e.g., ‘helps with depression,’ ‘relieves stress’) to imply that cannabis products are effective treatments for anxiety, depression, or other mental health conditions. This poses a serious risk to individuals seeking relief from symptoms of mental health conditions, as substantial evidence indicates that consuming Delta-9 tetrahydrocannabinol (primary intoxicating compound in cannabis) in large, acute doses or over prolonged periods can contribute to the onset of psychiatric conditions, worsen existing symptoms, and increase the risk of developing cannabis use disorder (Bahorik et al., 2018; Cuttler et al., 2018; Feingold et al., 2017; Hser et al., 2017; Kuhns et al., 2022; Moitra et al., 2021; National Academies of Sciences, Engineering, and Medicine et al., 2017; Patrick et al., 2016; Schoeler et al., 2018; van der Pol et al., 2013; Zvolensky et al., 2010). Future studies should examine how PACs shape purchasing behavior among individuals with mental health conditions and whether targeted mental health warning labels mitigate the effects of PACs.

This study has several limitations. First, participants were recruited through Meta® advertising, and more than half of the sample consisted of daily or near-daily cannabis consumers. As a result, the findings may not fully generalize to less frequent or inexperienced consumers, who may respond differently to the stimuli used in the store (Borodovsky, 2022; Borodovsky et al., 2018). Second, participants made hypothetical purchases. Although this study and others (Aston et al., 2021, 2015; Aston and Cassidy, 2019) provide evidence supporting the validity of hypothetical cannabis purchasing, future studies could incorporate designs that compare real-world in-person purchases with hypothetical POEMMS purchases to further examine the validity of spending behaviors. Third, participants provided highly variable realism ratings for the product prices. Because the sample was drawn from across the US, this variability may reflect differences in state-specific cannabis pricing and tax structures. Future research may benefit from using state-specific pricing models and restricting recruitment to a single state to improve perceived pricing realism.

In sum, this pilot study supports the use of POEMMS to examine online cannabis purchasing behavior. The platform realistically replicated key aspects of the consumer shopping experience and produced data consistent with real-world purchasing patterns. The findings lay a foundation for future research on the impact of online point-of-sale marketing strategies on cannabis consumer decisions and for regulatory efforts to protect public health.

## Data Availability

All data produced in the present study are available upon reasonable request to the authors

## Supported by

R21-DA062816, R01-DA050032, T32-DA037202, P30-DA037202

## Declaration of generative AI in scientific writing

Author JTB employed ChatGPT and Grammarly to help refine grammar, sentence structure, and word choice. The author(s) reviewed and retain full responsibility for the manuscript’s content.

## REFERENCES

Aston, E.R., Cassidy, R.N., 2019. Behavioral economic demand assessments in the addictions. Curr Opin Psychol 30, 42–47. 10.1016/j.copsyc.2019.01.016

Aston, E.R., Metrik, J., MacKillop, J., 2015. Further validation of a marijuana purchase task. Drug Alcohol Depend. 152, 32–38. 10.1016/j.drugalcdep.2015.04.025

Aston, E.R., Metrik, J., Rosen, R.K., Swift, R., MacKillop, J., 2021. Refining the marijuana purchase task: Using qualitative methods to inform measure development. Exp Clin Psychopharmacol 29, 23–35. 10.1037/pha0000355

Bahorik, A.L., Sterling, S.A., Campbell, C.I., Weisner, C., Ramo, D., Satre, D.D., 2018. Medical and non-medical marijuana use in depression: Longitudinal associations with suicidal ideation, everyday functioning, and psychiatry service utilization. J. Affect. Disord. 241, 8–14. 10.1016/j.jad.2018.05.065

Berg, C.J., LoParco, C.R., Cui, Y., Pannell, A., Kong, G., Griffith, L., Romm, K.F., Yang, Y.T., Wang, Y., Cavazos-Rehg, P.A., 2023a. A review of social media platform policies that address cannabis promotion, marketing and sales. Subst. Abuse Treat. Prev. Policy 18, 35. 10.1186/s13011-023-00546-x

Berg, C.J., Romm, K.F., Pannell, A., Sridharan, P., Sapra, T., Rajamahanty, A., Cui, Y., Wang, Y., Yang, Y.T., Cavazos-Rehg, P.A., 2023b. Cannabis retailer marketing strategies and regulatory compliance: A surveillance study of retailers in 5 US cities. Addict. Behav. 143, 107696. 10.1016/j.addbeh.2023.107696

Boatwright, K.D., Sperry, M.L., 2020. Accuracy of Medical Marijuana Claims Made by Popular Websites. J. Pharm. Pract. 33, 457–464. 10.1177/0897190018818907

Borodovsky, J.T., 2022. Generalizability and representativeness: Considerations for internet-based research on substance use behaviors. Exp. Clin. Psychopharmacol. 30, 466–477. 10.1037/pha0000581

Borodovsky, J.T., Budney, A.J., 2018. Cannabis regulatory science: risk-benefit considerations for mental disorders. Int Rev Psychiatry 30, 183–202. 10.1080/09540261.2018.1454406

Borodovsky, J.T., Marsch, L.A., Budney, A.J., 2018. Studying Cannabis Use Behaviors With Facebook and Web Surveys: Methods and Insights. JMIR Public Health Surveill 4, e48. 10.2196/publichealth.9408

Borodovsky, J.T., Sofis, M.J., Grucza, R.A., Budney, A.J., 2021. The importance of psychology for shaping legal cannabis regulation. Exp Clin Psychopharmacol 29, 99–115. 10.1037/pha0000362

Cao, Y., Carrillo, A.S., Zhu, S., Shi, Y., 2020. Point-of-Sale Marketing in Recreational Marijuana Dispensaries Around California Schools. J. Adolesc. Health 66, 72–78. 10.1016/j.jadohealth.2019.07.023

Cuttler, C., Spradlin, A., McLaughlin, R.J., 2018. A naturalistic examination of the perceived effects of cannabis on negative affect. J. Affect. Disord. 235, 198–205. 10.1016/j.jad.2018.04.054

D’Amico, E.J., Rodriguez, A., Dunbar, M.S., Firth, C.L., Tucker, J.S., Seelam, R., Pedersen, E.R., Davis, J.P., 2020. Sources of cannabis among young adults and associations with cannabis-related outcomes. Int. J. Drug Policy 86, 102971. 10.1016/j.drugpo.2020.102971

Donnan, J., Shogan, O., Bishop, L., Najafizada, M., 2022a. Drivers of purchase decisions for cannabis products among consumers in a legalized market: a qualitative study. BMC Public Health 22, 368. 10.1186/s12889-021-12399-9

Donnan, J., Shogan, O., Bishop, L., Swab, M., Najafizada, M., 2022b. Characteristics that influence purchase choice for cannabis products: a systematic review. J. Cannabis Res. 4, 9. 10.1186/s42238-022-00117-0

Donnan, J.R., Downey, M., Johnston, K., Najafizada, M., Bishop, L.D., 2024. Examining attributes of retailers that influence where cannabis is purchased: a discrete choice experiment. J. Cannabis Res. 6, 4. 10.1186/s42238-023-00204-w

Donnan, J.R., Johnston, K., Najafizada, M., Bishop, L.D., 2023. Drivers of Purchase Decisions Among Consumers of Dried Flower Cannabis Products: A Discrete Choice Experiment. J. Stud. Alcohol Drugs 84, 744–753. 10.15288/jsad.22-00269

Duan, Z., Kasson, E., Ruchelli, S., Rajamahanty, A., Williams, R., Sridharan, P., Sapra, T., Dopke, C., Pannell, A., Nakshatri, S., Berg, C.J., Cavazos-Rehg, P.A., 2023. Assessment of Online Marketing and Sales Practices Among Recreational Cannabis Retailers in Five U.S. Cities. Cannabis Cannabinoid Res. 10.1089/can.2022.0334

Feingold, D., Rehm, J., Lev-Ran, S., 2017. Cannabis use and the course and outcome of major depressive disorder: A population based longitudinal study. Psychiatry Res. 251, 225–234. 10.1016/j.psychres.2017.02.027

Gebru, N., Aston, E., Berey, B., Snell, L.M., Leeman, R., Metrik, J., 2023. “That’s Pot Culture Right There”: Purchasing Behaviors of People Who Use Cannabis Without a Medical Cannabis Card. Cannabis 6, 30–46.

Goodman, S., Leos-Toro, C., Hammond, D., 2019. The impact of plain packaging and health warnings on consumer appeal of cannabis products. Drug Alcohol Depend. 107633. 10.1016/j.drugalcdep.2019.107633

Hoeper, S., Crosbie, E., Holmes, L.M., Godoy, L., DeFrank, V., Hoang, C., Ling, P.M., 2022. “The Perfect Formula:” Evaluating Health Claims, Products and Pricing on Cannabis Dispensary Websites in Two Recently Legalized States. Subst. Use Misuse 57, 1207–1214. 10.1080/10826084.2022.2069267

Hser, Y.I., Mooney, L.J., Huang, D., Zhu, Y., Tomko, R.L., McClure, E., Chou, C.P., Gray, K.M., 2017. Reductions in cannabis use are associated with improvements in anxiety, depression, and sleep quality, but not quality of life. J Subst Abuse Treat 81, 53–58. 10.1016/j.jsat.2017.07.012

Israel, S., 2021. Online cannabis sales boom amid COVID-19 spurs delivery-service acquisitions. MJBizDaily. URL https://mjbizdaily.com/online-cannabis-sales-boom-amid-covid-19-spurs-delivery-service-acquisitions/ (accessed 9.1.22).

Jackson, M., 2020a. Marijuana shops cope with more online orders amid coronavirus pandemic. MJBizDaily. URL https://mjbizdaily.com/marijuana-shops-overhaul-workforces-to-cope-with-online-orders-curbside-delivery-boom-during-coronavirus-pandemic/ (accessed 9.4.22).

Jackson, M., 2020b. Live streaming, digital ads keep cannabis brands on consumers’ minds. MJBizDaily. URL https://mjbizdaily.com/live-streaming-digital-ads-help-cannabis-businesses-keep-brands-consumers-minds/ (accessed 9.4.22).

Kepple, N.J., Freisthler, B., 2018. Who’s Buying What and How Much? Correlates of Purchase Behaviors From Medical Marijuana Dispensaries in Los Angeles, California. J Prim Prev 39, 571–589. 10.1007/s10935-018-0528-5

Kepple, N.J., Mulholland, E., Freisthler, B., Schaper, E., 2016. Correlates of Amount Spent on Marijuana Buds During a Discrete Purchase at Medical Marijuana Dispensaries: Results from a Pilot Study. J Psychoact. Drugs 48, 50–5. 10.1080/02791072.2015.1116719

Kowitt, S.D., Yockey, R.A., Lee, J.G.L., Jarman, K.L., Gourdet, C.K., Ranney, L.M., 2022. The Impact of Cannabis Packaging Characteristics on Perceptions and Intentions. Am. J. Prev. Med. 63, 751–759. 10.1016/j.amepre.2022.04.030

Kuhns, L., Kroon, E., Colyer-Patel, K., Cousijn, J., 2022. Associations between cannabis use, cannabis use disorder, and mood disorders: longitudinal, genetic, and neurocognitive evidence. Psychopharmacology (Berl.) 239, 1231–1249. 10.1007/s00213-021-06001-8

Lau, N., Gerson, M., Korenstein, D., Keyhani, S., 2021. Internet Claims on the Health Benefits of Cannabis Use. J. Gen. Intern. Med. 36, 3611–3614. 10.1007/s11606-020-06421-w

Luc, M.H., Tsang, S.W., Thrul, J., Kennedy, R.D., Moran, M.B., 2020. Content analysis of online product descriptions from cannabis retailers in six US states. Int. J. Drug Policy 75, 102593. 10.1016/j.drugpo.2019.10.017

Marinello, S., Valek, R., Powell, L.M., 2024. Analysis of social media compliance with cannabis advertising regulations: evidence from recreational dispensaries in Illinois 1-year post-legalization. J. Cannabis Res. 6, 2. 10.1186/s42238-023-00208-6

McCann, M., 2024. Cannabis Consumers in America 2023: Part 3. New Frontier Data.

Moitra, E., Anderson, B.J., Herman, D.S., Stein, M.D., 2021. Longitudinal examination of coping-motivated marijuana use and problematic outcomes among emerging adults. Addict. Behav. 113, 106691. 10.1016/j.addbeh.2020.106691

Mutti-Packer, S., Collyer, B., Hodgins, D.C., 2018. Perceptions of plain packaging and health warning labels for cannabis among young adults: findings from an experimental study. BMC Public Health 18, 1361. 10.1186/s12889-018-6247-2

National Academies of Sciences, Engineering, and Medicine, Health and Medicine Division, Board on Population Health and Public Health Practice, Committee on the Health Effects of Marijuana: An Evidence Review and Research Agenda, 2017. The Health Effects of Cannabis and Cannabinoids: The Current State of Evidence and Recommendations for Research, The National Academies Collection: Reports funded by National Institutes of Health. National Academies Press (US), Washington (DC).

Olsen, M.C., Smith, K.M., 2019. The cannabis industry: a natural laboratory for marketing strategy research. Mark. Lett. 31, 7–12. 10.1007/s11002-019-09502-x

Olson, B., 2015. Chart of the Week: Average Purchase Amount at Dispensaries Ranges From $60 to $100+. MJBizDaily. URL https://mjbizdaily.com/chart-week-average-marijuana-dispensary-purchase-amounts-range-60-100/ (accessed 7.31.23).

Patrick, M.E., Bray, B.C., Berglund, P.A., 2016. Reasons for Marijuana Use Among Young Adults and Long-Term Associations With Marijuana Use and Problems. J. Stud. Alcohol Drugs 77, 881–888. 10.15288/jsad.2016.77.881

Rhee, S., Jung, W., 2023. “There must be something good”: Fair balance and ad appeal of marijuana brands’ website. Int. J. Drug Policy 119, 104116. 10.1016/j.drugpo.2023.104116

Rup, J., Goodman, S., Hammond, D., 2020. Cannabis advertising, promotion and branding: Differences in consumer exposure between ‘legal’ and ‘illegal’ markets in Canada and the US. Prev. Med. 133, 106013. 10.1016/j.ypmed.2020.106013

Schoeler, T., Theobald, D., Pingault, J.-B., Farrington, D.P., Coid, J.W., Bhattacharyya, S., 2018. Developmental sensitivity to cannabis use patterns and risk for major depressive disorder in mid-life: findings from 40 years of follow-up. Psychol. Med. 48, 2169–2176. 10.1017/S0033291717003658

Shi, Y., Cao, Y., Shang, C., Pacula, R.L., 2019. The impacts of potency, warning messages, and price on preferences for Cannabis flower products. Int J Drug Policy 74, 1–10. 10.1016/j.drugpo.2019.07.037

Statista, 2020. Average spend per cannabis purchase U.S. 2020 [WWW Document]. Statista. URL https://www.statista.com/statistics/1281579/us-average-cannabis-spend-per-purchase/ (accessed 7.31.23).

Team Headset, 2020. What does the average cannabis consumer look like? [WWW Document]. URL https://www.headset.io/industry-reports/what-does-the-average-cannabis-consumer-look-like (accessed 3.12.25).

Trangenstein, P.J., Whitehill, J.M., Jenkins, M.C., Jernigan, D.H., Moreno, M.A., 2021. Cannabis Marketing and Problematic Cannabis Use Among Adolescents. J. Stud. Alcohol Drugs 82, 288–296. 10.15288/jsad.2021.82.288

van der Pol, P., Liebregts, N., de Graaf, R., Korf, D.J., van den Brink, W., van Laar, M., 2013. Predicting the transition from frequent cannabis use to cannabis dependence: a three-year prospective study. Drug Alcohol Depend 133, 352–9. 10.1016/j.drugalcdep.2013.06.009

Whitehill, J.M., Trangenstein, P.J., Jenkins, M.C., Jernigan, D.H., Moreno, M.A., 2020. Exposure to Cannabis Marketing in Social and Traditional Media and Past-Year Use Among Adolescents in States With Legal Retail Cannabis. J Adolesc Health 66, 247–254. 10.1016/j.jadohealth.2019.08.024

Zhu, B., Guo, H., Cao, Y., An, R., Shi, Y., 2021. Perceived Importance of Factors in Cannabis Purchase Decisions: A Best-worst Scaling Experiment. Int. J. Drug Policy 91, 102793. 10.1016/j.drugpo.2020.102793

Zvolensky, M.J., Cougle, J.R., Johnson, K.A., Bonn-Miller, M.O., Bernstein, A., 2010. Marijuana use and panic psychopathology among a representative sample of adults. Exp. Clin. Psychopharmacol. 18, 129–134. 10.1037/a0019022

